# Investigating the relationship between DNA methylation, genetic variation, and suicide attempt in bipolar disorder

**DOI:** 10.1101/2024.04.03.24305263

**Authors:** Aysheh Alrfooh, Lucas G. Casten, Jenny Gringer Richards, John A. Wemmie, Vincent A. Magnotta, Jess G. Fiedorowicz, Jacob Michaelson, Aislinn J. Williams, Marie E. Gaine

## Abstract

Individuals with bipolar disorder are at increased risk for suicide, and this can be influenced by a range of biological, clinical, and environmental risk factors. Biological components associated with suicide include DNA modifications that lead to changes in gene expression. Common genetic variation and DNA methylation changes are some of the most frequent types of DNA findings associated with an increased risk for suicidal behavior. Importantly, the interplay between genetic predisposition and DNA methylation patterns is becoming more prevalent in genetic studies. We hypothesized that DNA methylation patterns in specific loci already genetically associated with suicide would be altered in individuals with bipolar disorder and a history of suicide attempt. To test this hypothesis, we searched the literature to identify common genetic variants (N=34) previously associated with suicidal thoughts and behaviors in individuals with bipolar disorder. We then created a customized sequencing panel that covered our chosen genomic loci. We profiled DNA methylation patterns from blood samples collected from bipolar disorder participants with suicidal behavior (N=55) and without suicidal behavior (N=51). We identified seven differentially methylated CpG sites and five differentially methylated regions between the two groups. Additionally, we found that DNA methylation changes in *MIF* and *CACNA1C* were associated with lethality or number of suicide attempts. Finally, we identified three meQTLs in *SIRT1*, *IMPA2*, and *INPP1*. This study illustrates that DNA methylation is altered in individuals with bipolar disorder and a history of suicide attempts in regions known to harbor suicide-related variants.

## Introduction

Bipolar disorder (BD) is a lifelong, classically episodic, illness and has the highest risk for suicide of all mental disorders. The annual risk for suicide attempts in individuals with BD has been estimated to be 30-60 fold higher than the general population [1]. Suicide in BD is also multi-factorial, with numerous biological, clinical, and environmental factors known to increase risk for suicide-related outcomes. Biological risk factors for suicidal behavior include genetic and epigenetic variation, which has been researched in varying degrees in BD.

The hypothesis that attempted suicide in BD has a genetic component has been established for several years. Genome-wide association studies (GWAS) have identified several risk loci associated with suicidal behavior in BD [1–7], mainly pointing towards pathways such as serotonin signaling and the hypothalamic-pituitary-adrenal (HPA) axis. Notably, some studies show that there may be distinct genetic risk factors, such as Acid Phosphatase 1 (*ACP1*), associated with suicide in individuals with BD specifically [2]. One of the largest GWAS to focus on BD separately found one significant locus within the BD cohort on chromosome 4 in *LOC105374524* [3]. However, the modest effect sizes suggest additional factors may need to be considered.

Unlike genetic variation, DNA methylation can be dynamic and change throughout our lifetime. DNA methylation patterns link our genes and our environment by altering gene expression in the presence of environmental factors. Significant differences in DNA methylation have been observed between those with BD with and without a history of suicidal behavior. Individuals with BD and a high suicidality score showed hypermethylation at Nucleoporin 133 (*NUP133*), hypomethylation at MAGUK P55 Scaffold Protein 4 (*MPP4*), and hypomethylation at TBC1 Domain Family Member 16 (*TBC1D16*) [8]. A study of those with a history of suicide attempts and BD found several differentially methylated CpG sites and regions with the strongest findings located in C-X-C Motif Chemokine Ligand 8 (*CXCL8*) and Tripartite Motif Containing 40 (*TRIM40*), both inflammatory genes [9]. A targeted sequencing study focused on suicide death investigated over 3.7 million CpG sites and reported hypomethylation at Rho Guanine Nucleotide Exchange Factor 38 (*ARHGEF38)* [10]. More recently, studies have shown that epigenetic aging is increased in individuals with BD and a history of suicide attempts [11]. Despite interesting and biologically relevant findings, many DNA methylation changes associated with suicide in BD have yet to be replicated in independent cohorts.

One component that could be causing variability across methylome studies is the disruption of certain DNA methylation patterns by genetic variation. This can contribute to inter-individual differences and lead to changes in gene expression and phenotypic traits. Regions of the genome that harbor genetic variants capable of influencing DNA methylation patterns are called methylation quantitative trait loci (meQTLs) [12–16]. Notably, the first suicide study to investigate DNA methylation did it in the context of a genetic variant [17]. They studied allele-specific DNA methylation associated with the Serotonin Receptor 2A (*HTR2A*) C102T locus in individuals who died by suicide and found associations with schizophrenia but not BD. Another study found that a genetic variant in the Spindle and Kinetochore Associated Complex Subunit 2 (*SKA2*) gene influenced an association between DNA methylation and suicide death [18]. A recent study identified three meQTLs in Mitotic Arrest Deficient 1 Like 1 (*MAD1L1*) associated with suicide severity and depression risk [19]. These studies indicate that genetic variation should be taken into account when interpreting methylome analyses of suicidal behavior.

In this study, we hypothesized that loci harboring genetic variants associated with suicide in BD would contain differentially methylated CpG sites and regions increasing the risk for suicidal behavior. We additionally hypothesized that genetic variants in these regions would influence DNA methylation patterns as meQTLs. To test these hypotheses, we performed a targeted bisulfite sequencing study to quantify DNA methylation and identify meQTLs in suicide-related loci, which were chosen based on a literature review of genetic studies investigating suicidal thoughts and behaviors in BD.

## Material and Methods

### Participants and Sample Collection

The Iowa Neuroscience Institute’s Bipolar Disorder Research Program of Excellence (BD-RPOE) recruited participants, after University of Iowa Institutional Review Board approval, for this study as previously described [9, 11, 20–28]. Written informed consent was provided by all participants and most also provided a blood sample for epi/genetic analyses. The subjects in the BD group ranged in age from 18 to 70 years old, had the ability to consent, and were diagnosed with BD Type I according to the Structured Clinical Interview for DSM Disorders (SCID). The Columbia Suicide Severity Rating Scale was used to record suicide history [29]. The variables used in this study were history of suicide attempts, total number of attempts in their lifetime, and severity of most lethal attempt. For our study, we included a total of 160 samples (from 106 BD participants, 55 participants with a history of suicide attempt, and 54 non-BD controls; **Supplemental Table 1**).

### Study Design

We aimed to study the correlation between DNA methylation patterns in genomic regions previously associated with suicidal behavior in BD. To do this, we completed a literature review to identify common genetic variants that showed an association with suicidal thoughts and behaviors in BD. In total, we found 40 common genetic variants that showed a significant association with suicidal thoughts and behaviors in BD. We included genetic variants with nominal significance because *cis*-meQTLs are enriched for low GWAS P-values in a variety of phenotypes [13]. In our final design, we only included 34 genetic variants (**Table 1**) due to the technical difficulties in designing primers that covered both genetic variants and CpG sites in bisulfite-converted DNA in three of the original 40 loci. The chosen loci contained 106 CpG sites for further investigation.

**Table 1.**
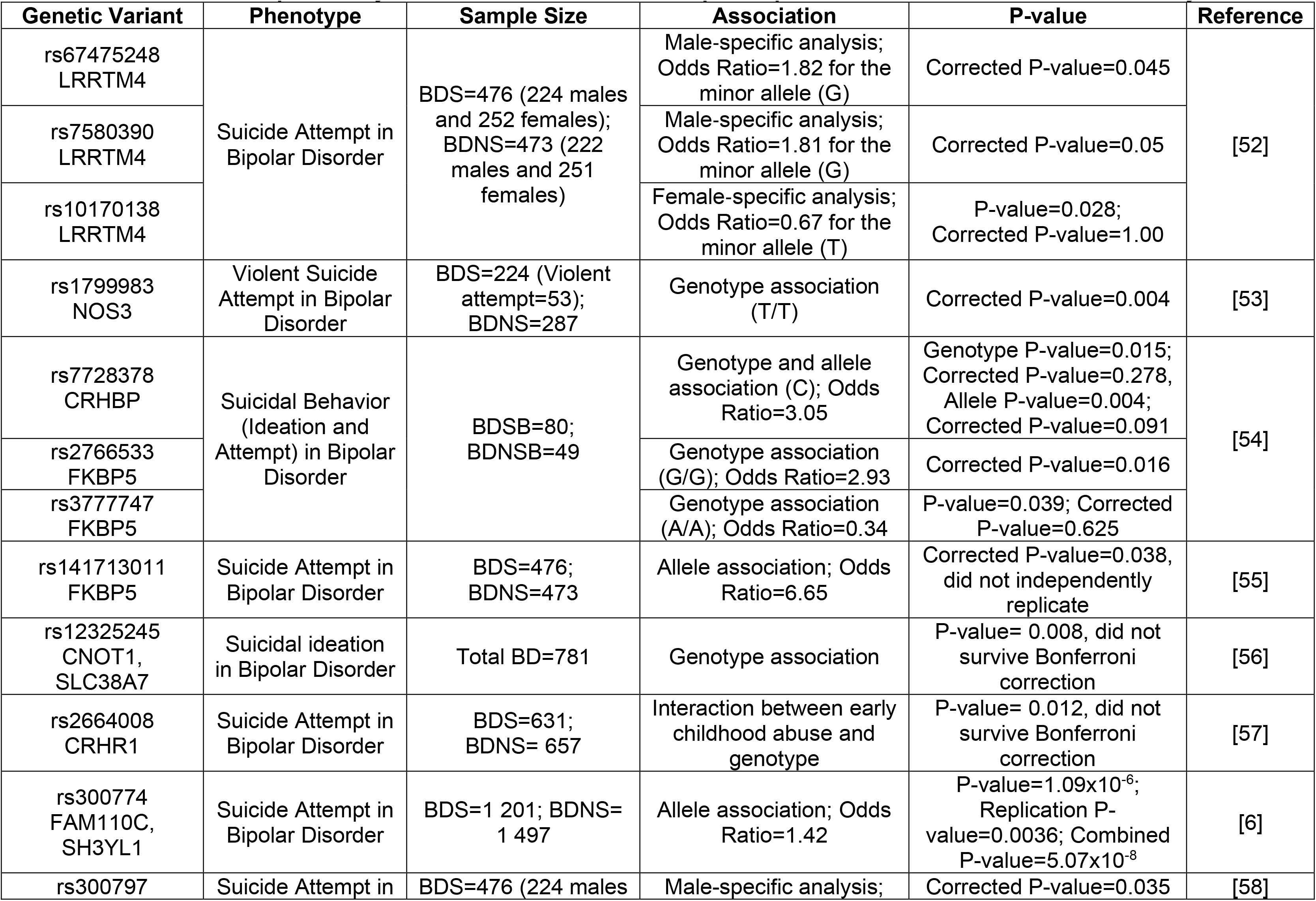
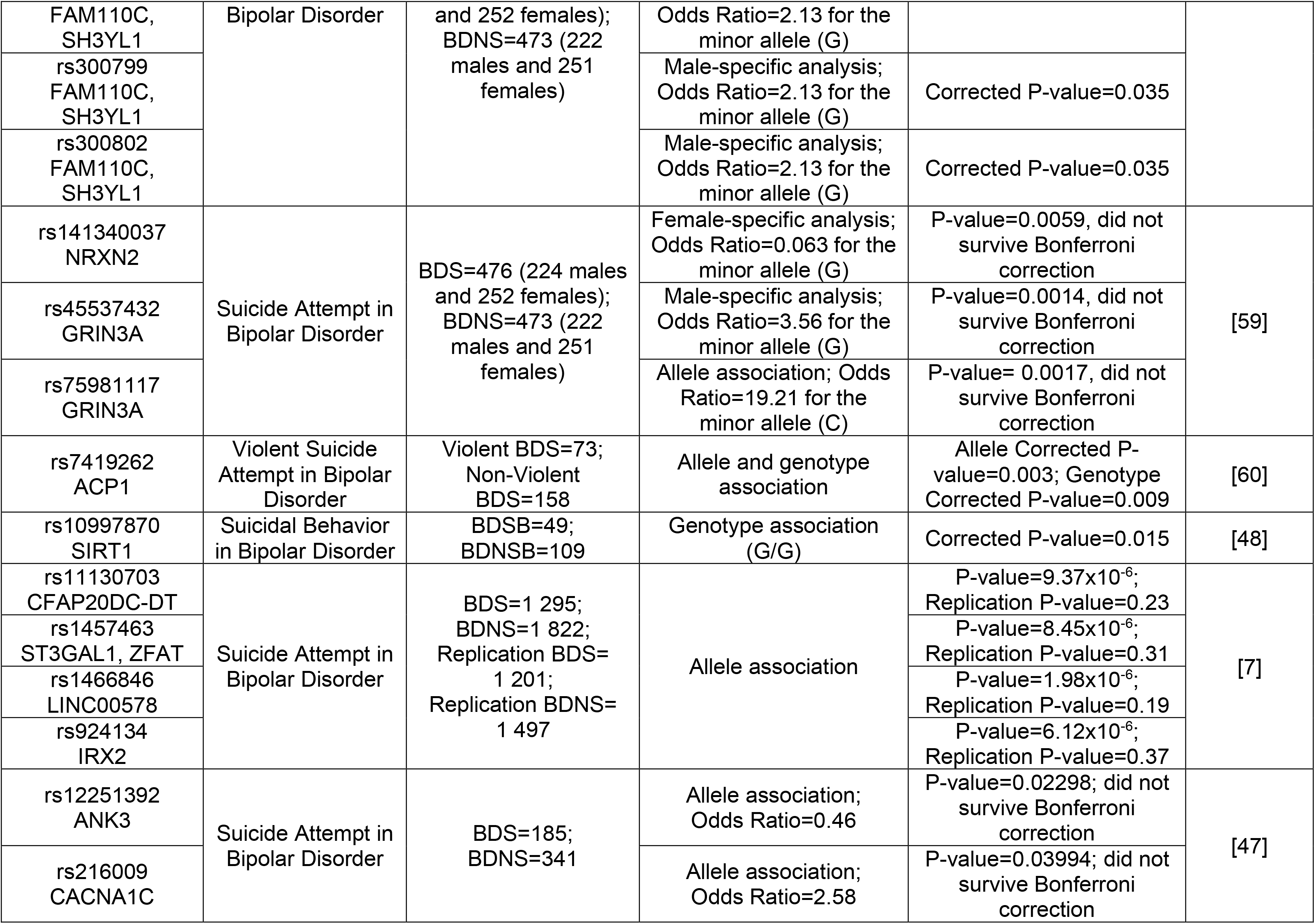
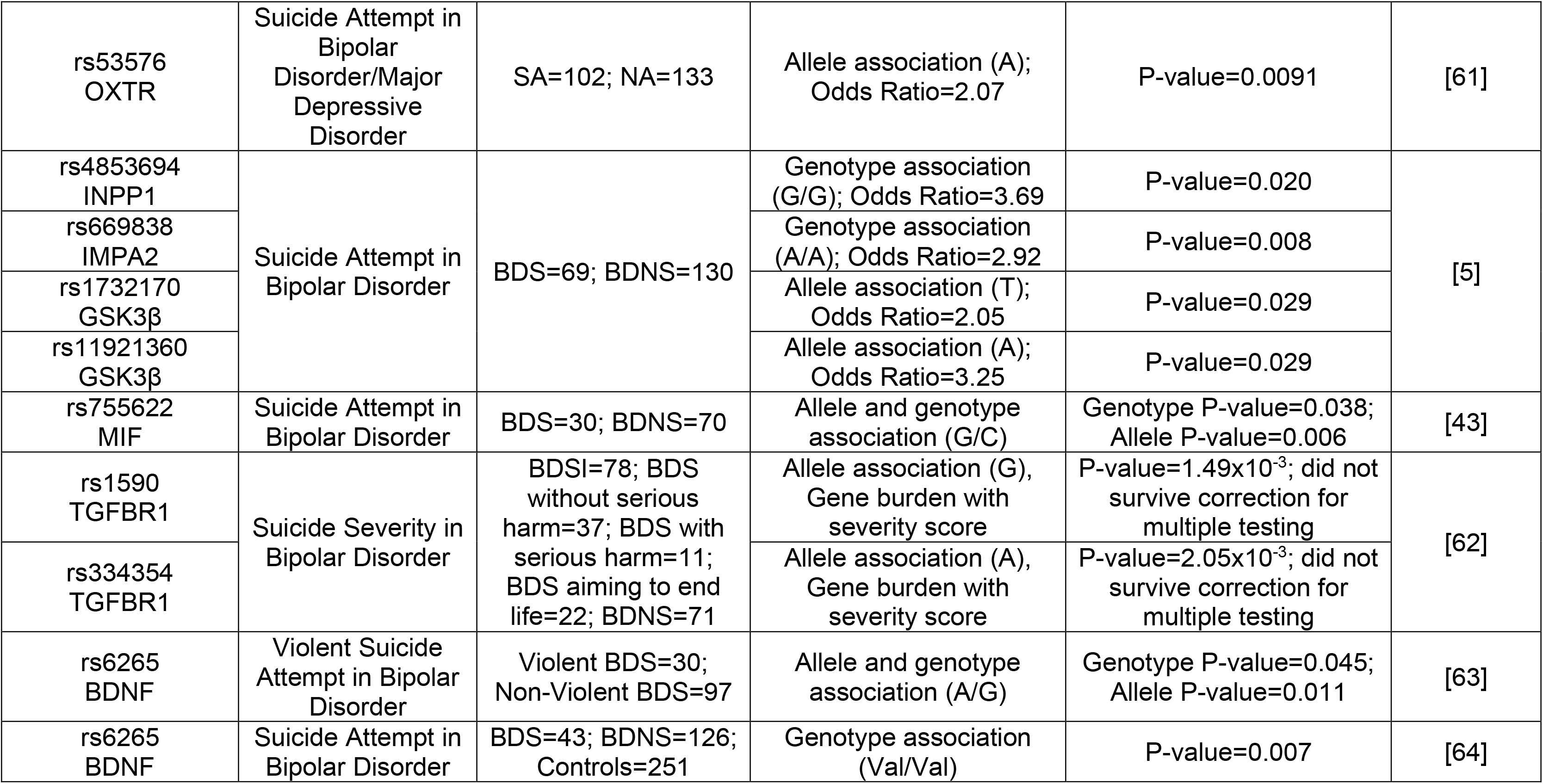
Genetic variants previously associated with suicide attempt in bipolar disorder and included in this study. A literature review was conducted to identify common genetic variants associated with suicidal thoughts and behaviors in bipolar disorder. Only variants included in the final analysis are described in the table (N=34). BD, Bipolar Disorder. BDS, individual with bipolar disorder and a history of suicide attempts. BDNS, individual with bipolar disorder and no history of suicide attempts. BDSB, individual with bipolar disorder and a history of suicidal behavior. BDNSB, individual with bipolar disorder and no history of suicidal behavior. BDSI, individual with bipolar disorder and a history of suicidal ideation. SA, individual with a history of suicide attempts. NA, individual with no history of suicide attempts.

We completed four analyses to identify differentially methylated CpG sites, differentially methylated regions, correlations between the number of suicide attempts and lethality of suicide attempts, and meQTLs. We employed a case-only approach and removed any findings that were also significantly different when comparing BD subjects (with and without a history of suicide attempts) to controls to identify only suicide-specific findings.

### Sample Preparation and Sequencing

Extracted DNA (100 ng) from whole blood was bisulfite converted using MethylCode Bisulfite Conversion Kit following the protocol guide. Sequencing libraries were prepared using the Ion AmpliSeq™ Library Kit 2.0 and Ion Xpress™ Barcode Adapters 1-16 Kit according to manufacturer instructions with some modifications. The samples were processed in 26 separate runs, with each run comprising 8-10 samples, depending on the inclusion of high-quality DNA quality controls (Jurkat Genomic DNA, ThermoFisher).

### Bioinformatics Pipeline

Before base calling, the Ion Torrent system eliminated polyclonal reads, “low quality” reads based on the data noise, and reads that were too short (less than 25 bp). Base calling was followed by quality trimming to filter all bases that had low-quality scores less than 15 and to trim the 5’ end using a 30 bp window. Raw data files were processed using the methylation_analysis plugin/pipeline (provided by S5 Torrent Server VM). Eleven samples were excluded because of low-quality run metrics or poor conversion rate (<93% or NA). Technical replicates were utilized across runs to increase the mean depth coverage of samples with less than 2,000 reads. In total 149 unique samples remained after filtering and combining technical replicates.

Downstream analyses of the methylation call files were completed using the R package, methylKit [30]. CpG sites with low coverage (less than 10 reads) were removed, and reads were normalized using the median read coverage as a scaling factor in methylKit. Any CpG site that had more than 99.9% coverage in each sample was filtered and both strands merged. To adjust for technical variation, we conducted a principal component analysis (PCA) and removed the first and second principal components from the methylation base objects. After filtering, 88 CpG sites remained for analysis.

### Differential Methylation Analysis

Using methylKit, we investigated differential DNA methylation at specific CpG sites. We employed a logistic regression model where age, sex, race, body mass index (BMI), and smoking history were included as covariates. Because DNA methylation data variance (errors or residuals) was more variable than expected, we corrected for overdispersion. We also investigated differentially methylated regions (windows of 300 bp step-size). The sliding linear model method (SLIM, in methylKit) was used to adjust for multiple comparisons [31]. In order to identify biological changes associated with suicide only, we also compared BD subjects (with and without a history of suicide attempts) to controls and removed any findings that were also significantly associated with BD. By employing this filtering method, we were able to focus on DNA methylation changes associated only with suicide.

### Correlation Analyses

As an exploratory analysis, we investigated the relationship between DNA methylation patterns and the frequency of suicide attempts or severity of attempt to determine if there was a cumulative association. We tested the correlation between the number of lifetime suicide attempts with DNA methylation percentage at 88 CpG sites in participants with BD and a history of suicide attempts (N=49) using the cor.test () function in R. Since both DNA methylation percentage and the number of lifetime suicide attempts were not normally distributed, a non-parametric test (Kendall’s test) was used to measure the monotonic relationship between the two variables. To test the correlation between severity of attempt and DNA methylation percentage, the same function was used. Extreme (respiratory arrest, coma), severe (throat cut), moderate (briefly unconscious), mild (took pills, upset stomach), minimum (no effect), and no danger were assigned a numerical value for the analysis. Due to the multiple testing burden, False Discovery Rate (FDR) p-value adjustment was used to correct for multiple comparisons.

### Genotyping and Imputation

We were unable to use the bisulfited sequence reads to confidently call genotypes in our samples. However, samples in the same cohort were genotyped on Illumina’s Infinium PsychArray-24 v1.3 BeadChip using GenomeStudio 2.0 software with the cluster files provided by Illumina (N=112). Genotype data was exported to PLINK binary format for quality control and imputation [32]. Next, we filtered genetic variants by removing indels and non-biallelic sites. Variants were removed if they had low minor allele frequency in our sample (<5%) or if they defied Hardy-Weinberg equilibrium (P-value<1×10^−6^). All samples had high genotyping rates (>99%). Individuals were removed due to relatedness (identity-by-descent cutoff of 0.125), high heterozygosity rates (removed when using the PLINK “--het” command), or if they did not cluster with the European superpopulation in 1000 Genomes (to reduce effects of population stratification in subsequent analysis) [33]. A total of 102 samples and 350,345 common variants passed all quality control measures and were used for imputation and downstream analysis.

Data passing quality control were then imputed to the HRC r1.1 hg19 build reference dataset [34] using Michigan Imputation Server MiniMac4 [35]. Variants with imputation R-squared scores less than 0.3 were removed (final variant N=12,507,615). A cutoff of 90% probability was used to convert imputed genotype dosages to hard calls. Finally, to identify ancestrally relevant genetic principal components, we conducted a PCA in 1000 Genomes Europeans using HapMap3 genetic variants that were also present in our dataset with the bigsnpr R package [36]. We then projected our samples onto the 1000 Genomes PCA dimensions.

### meQTL Analysis

The R package, MatrixEQTL, was used to identify meQTLs [37]. Data transformation of the methylation percentage data was required because it was not normally distributed. We focused our analysis on common variants (minor allele frequency >5%) within 15Kb of the CpG sites (*cis*-meQTLs; based on hg19 coordinates of CpG sites and variant genotypes) [38]. A total of 864 variants from the imputed genotype data were within the regions of interest and were analyzed (mapping nearby 55 of the CpG sites). We included age, sex, BMI, smoking history, BD diagnosis and the first five genetic principal components as covariates. A total of 3,516 meQTL tests were conducted. Due to the multiple testing burden, FDR p-value adjustment was used to correct for multiple comparisons.

## Results

### Demographic information

Detailed clinical and demographic data on the cohort used are shown in **Supplemental Table 1**. The BD cohort was separated into individuals with a history of suicide attempt (N=55) and individuals with no history of suicide attempt (N=51). These groups were not significantly different in sex, age, race, or BMI (P-value>0.05). Individuals with a history of suicide attempt did have significantly more smoking history (P-value=0.024) and higher numbers of lifetime attempts (P-value=1.83×10^−6^).

### DNA methylation patterns in loci genetically associated with attempted suicide in BD

Our primary analysis was to investigate the DNA methylation status of 88 CpG sites adjacent to 34 genetic variants previously associated with suicidal thoughts and behaviors. We identified seven CpG sites differentially methylated in individuals with BD and a history of suicide attempt. The identified CpG sites were located close to six genes previously linked to suicidal behavior (**Table 2**). The most significant finding was a CpG site located in the 3’UTR of Transforming Growth Factor Beta Receptor 1 (*TGFBR1*; Q-value=2.96×10^−4^).

**Table 2.** Distinct differentially methylated CpG sites associated only with a history of suicide attempts. Results with a q-value less than 0.05 are shown. Findings were filtered against bipolar disorder vs. controls and bipolar disorder with a history of suicide attempts vs. controls analyses to include suicide-specific results. GRCh38/hg38 used to annotate genomic locations.

| **Gene** | **Nearby Suicide-Related Variant** | **CpG Genomic Location** | **P-value** | **Q-value** | **Methylation Difference** |
| --- | --- | --- | --- | --- | --- |
| TGFBR1 | rs1590 (chr9:99153883) | chr9:99153803 | $4.35 \times 10^{-6}$ | $2.96 \times 10^{-4}$ | -26.350 |
| MIF | rs755622 (chr22:23894205) | chr22:23894167 | $1.89 \times 10^{-4}$ | $4.29 \times 10^{-3}$ | -3.552 |
| GSK3β | rs1732170 (chr3:119864829) | chr3:119864633 | $4.44 \times 10^{-4}$ | $6.05 \times 10^{-3}$ | -3.938 |
| NOS3 | rs1799983 (chr7:150999023) | chr7:150999085 | $4.02 \times 10^{-3}$ | 0.034 | 7.712 |
| BDNF | rs6265 (chr11:27658369) | chr11:27658375 | $5.88 \times 10^{-3}$ | 0.036 | 7.896 |
| FKBP5 | rs2766533 (chr6:35717713) | chr6:35717775 | $4.95 \times 10^{-3}$ | 0.036 | 1.208 |
| FKBP5 | rs3777747 (chr6:35611225) | chr6:35611420 | $9.11 \times 10^{-3}$ | 0.048 | -8.756 |

We also identified five differentially methylated regions in individuals with BD and a history of suicide attempt (**Table 3**). These regions showed multiple CpG sites (between 2-6 CpG sites) with altered DNA methylation patterns associated with suicide (most significant Q-value=1.63×10^−7^). Three of these regions contained a differentially methylated CpG site identified in the previous single CpG site analysis.

**Table 3.** Distinct differentially methylated regions associated only with a history of suicide attempts. Results with a q-value less than 0.05 are shown. Only common genetic variants (MAF>0.05) were included. CpG sites highlighted green also showed a significant difference in DNA methylation at the single CpG level. Findings were filtered against bipolar disorder vs. controls and bipolar disorder with a history of suicide attempts vs. controls analyses to include suicide-specific results. GRCh38/hg38 used to annotate genomic locations.

| Gene | Nearby Suicide-Related Variant | Genomic Location | P-value | Q-value | Methylation Difference | Number of CpG Sites in the Region | CpG Site Genomic Location |
| --- | --- | --- | --- | --- | --- | --- | --- |
| TGFB1 | rs1590 (chr9:99153883, exon) | chr9:99153601-99153900 | 1.66x10<sup>-8</sup> | 1.63x10<sup>-7</sup> | -11.918 | 2 | chr9:99153797<br>chr9:99153803 |
| BDNF | rs6265 (chr11:27658369, exon) | chr11:27658201-27658500 | 5.57x10<sup>-3</sup> | 0.018 | 1.407 | 6 | chr11:27658292<br>chr11:27658306<br>chr11:27658332<br>chr11:27658375<br>chr11:27658429<br>chr11:27658452 |
| FKBP5 | rs3777747 (chr6:35611225, intron) | chr6:35611201-35611500 | 0.010 | 0.025 | -8.253 | 2 | chr6:35611312<br>chr6:35611420 |
| LRRTM4 | rs67475248 (chr2:76864739, intron) | chr2:76864801-76865100 | 0.025 | 0.030 | 10.346 | 2 | chr2:76864819<br>chr2:76864851 |
| INPP1 | rs4853694 (chr2:190354547, intron) | chr2:190354501-190354800 | 0.023 | 0.030 | 2.255 | 4 | chr2:190354502<br>chr2:190354539<br>chr2:190354545<br>chr2:190354582 |

### Correlation between DNA methylation and number of suicide attempts

We completed an exploratory analysis to investigate if DNA methylation patterns in suicide-related loci correlated with the number of suicide attempts. From the 88 CpG sites that were included, 16 CpG sites were correlated (P-value<0.05) with the number of lifetime suicide attempts (**Supplemental Table 2**). The majority of the CpG sites (N=13) showed a negative correlation with the number of lifetime suicide attempts. Only one CpG site (chr22:23894125), located just upstream of Macrophage Inhibitory Factor (*MIF*), survived correction for multiple testing (FDR=0.022; **Table 4**). For this CpG site, hypomethylation was correlated with an increased number of suicide attempts.

**Table 4.**
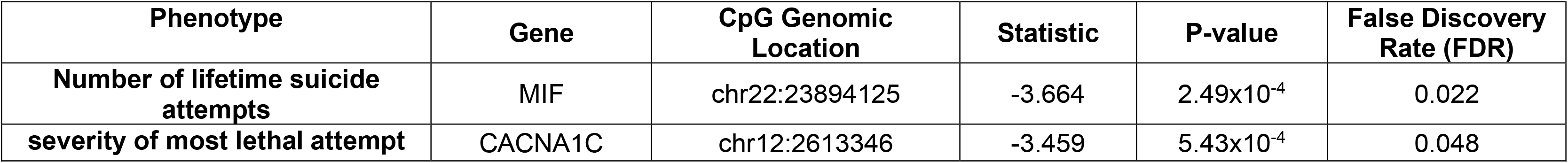
Correlation between DNA methylation, number of lifetime suicide attempts, and severity of most lethal attempt. Results with a False Discovery Rate (FDR) less than 0.05 are shown. GRCh38/hg38 used to annotate genomic locations.

| **Phenotype** | **Gene** | **CpG Genomic Location** | **Statistic** | **P-value** | **False Discovery Rate (FDR)** |
| --- | --- | --- | --- | --- | --- |
| **Number of lifetime suicide attempts** | MIF | chr22:23894125 | -3.664 | $2.49 \times 10^{-4}$ | 0.022 |
| **severity of most lethal attempt** | CACNA1C | chr12:2613346 | -3.459 | $5.43 \times 10^{-4}$ | 0.048 |

### Correlation between DNA methylation and lethality of attempt

We completed another exploratory analysis to investigate if DNA methylation patterns in suicide-related loci correlated with the severity of the most lethal attempt. From the 88 CpG sites that were included, seven CpG sites showed a correlation (P-value<0.05) with the lethality of attempt (**Supplemental Table 3**). The severity of the most lethal attempt showed a negative correlation with DNA methylation at six CpG sites and a positive correlation with one CpG site. CpG site chr12:2613346, which is located in an intron of Calcium Voltage-Gated Channel Subunit Alpha1 C (*CACNA1C*), was the only CpG site to survive correction for multiple testing (FDR=0.048; **Table 4**). For this CpG site, DNA methylation was negatively correlated with severity, with hypomethylation associated with extreme severity (**Figure 1**).

**Figure 1.**
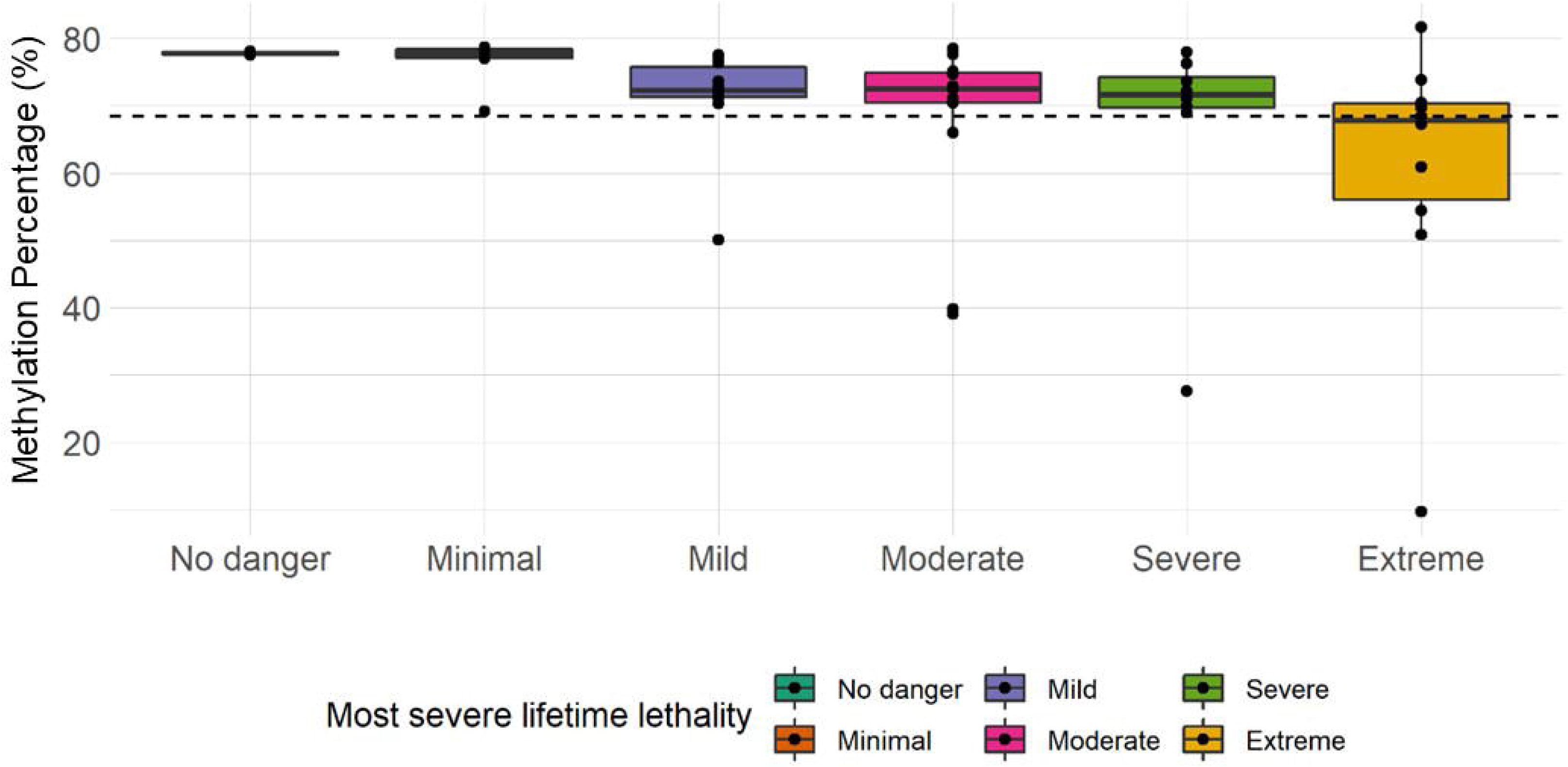
Methylation of a *CACNA1C* CpG site negatively correlated with severity of suicide attempt. A CpG site located at chr12:2613346, which lies in intron 29 of *CACNA1C*, was negatively correlated with severity of most lethal suicide attempt. False discovery rate (FDR) was less than 0.05. The dotted line represents the mean.

### Suicide-related loci harbor meQTLs

We hypothesized that, within our target regions, certain genetic variation would be associated with DNA methylation patterns. To test this, we investigated *cis*-meQTLs in our data including 55 CpG sites and 864 genetic variants. In total, 3 *cis*-meQTL pairs survived correction for multiple testing (FDR<0.05; **Table 5**). Three CpG sites were enriched in these meQTLs (chr10:67908277, chr18:11994474, and chr2:190354545) located in three suicide-related genes (Sirtuin 1 (*SIRT1*), Inositol monophosphatase 2 (*IMPA2*), and Inositol polyphosphate-1-phosphatase (*INPP1*)). All the significant meQTLs were associated with hypomethylation.

**Table 5.** *Cis*-meQTL analysis results. Results with a False Discovery Rate (FDR) less than 0.05 are shown. GRCh38/hg38 used to annotate genomic locations.

| **Gene** | **CpG site Location** | **Genetic Variant** | **Base Pair Distance** | **Effect Allele** | **Other Allele** | **Effect Allele Frequency** | **Beta** | **Statistic** | **P-value** | **False Discovery Rate (FDR)** |
| --- | --- | --- | --- | --- | --- | --- | --- | --- | --- | --- |
| SIRT1 | chr10:<br>67908277 | rs2234975 | -10044 | T | C | 0.054 | -0.191 | -4.914 | 3.98E-06 | 0.014 |
| IMPA2 | chr18:<br>11994474 | rs57967783 | 10231 | T | C | 0.070 | -0.440 | -4.610 | 1.32E-05 | 0.023 |
| INPP1 | chr2:<br>190354545 | rs2067390 | 10243 | A | T | 0.069 | -0.136 | -4.353 | 3.55E-05 | 0.042 |

## Discussion

The field of suicide epigenetics, across disorders and in BD, has grown substantially in the past decade. However, few studies have been replicated, and this may partly be due to the potential interplay between genetic predisposition and DNA methylation associated with suicide. Using our well-phenotyped cohort, we report differentially methylated patterns located close to genetic loci that have previously been associated with suicidal thoughts and behavior in BD. Several suicide-related gene loci were found to harbor significantly differentially methylated CpG sites and/or regions associated with suicide attempt in BD. Two additional CpG sites were associated with number and severity of attempts in the *MIF* and *CACNA1C* genes respectively. Our meQTL analysis also yielded significant findings in *SIRT1*, *IMPA2*, and *INPP1*.

Using a targeted sequencing approach, we were able to identify significant DNA methylation changes in genomic regions previously associated with suicide. These changes in eight genes suggest that they are biologically relevant to suicide in BD, either genetically, epigenetically, or both. Notably, six of these genes are related to immune response. Other studies have identified immune abnormalities associated with suicidal behavior, including altered inflammatory markers and links to autoimmune disorders [39–41]. Our work is also consistent with a recent epigenetic study focused on suicide attempt in BD that found altered DNA methylation in immune-related genes [9]. It is difficult to separate whether these DNA methylation changes are a causal factor of suicide or a consequence of the suicide attempt.

However, our work also highlighted a more nuanced relationship between DNA methylation and suicide severity by investigating number of suicide attempts and severity of most lethal attempt. We found that hypomethylation of a CpG site in *MIF* was correlated with number of suicide attempts. *MIF* is an inflammatory mediator that is released by a variety of immune cells in response to stimuli such as hypoxia, and studies of *MIF* propose that it is associated with innate immune response [42]. Genetic variation in *MIF* has been associated with suicide attempt in BD [43], but this was not replicated in a Japanese population [44]. This correlative relationship highlights the possibility that altered DNA methylation of certain immune-related genes may be a consequence of the stress of suicide attempts.

Hypomethylation of a CpG site in *CACNA1C* was correlated with severity of attempt. *CACNA1C* regulates neuronal plasticity and is a well-known risk gene for neuropsychiatric disorders including BD [45]. An imaging study of individuals with a history of suicide attempt found that *CACNA1C* DNA methylation was associated with altered brain activity [46]. A genetic variant in *CACNA1C* was associated with suicide attempt in only BD, but this did not survive correction for multiple testing [47]. This finding is consistent with *MIF*, highlighting that altered DNA methylation may be a consequence of the severity of the attempt. It is also possible that severe suicide attempts represent a subtype of suicide with distinct biological mechanisms. It is notable that our data suggests specific genes are vulnerable to these stressors as not all genomic loci studied were impacted. Overall, epigenetic changes related to immune response are prevalent in blood samples from individuals with BD and a history of suicide attempts, but longitudinal studies are required to fully uncover how suicide attempts change DNA methylation.

The presence of significant meQTLs in our target loci suggests that suicide-related genes harbor genetic variants that influence DNA methylation. The significant meQTLs were located in *SIRT1*, *IMPA2*, and *INPP1*. Genetic variation in *SIRT1* has been linked to suicide risk in BD [48] and major depressive disorder [49]. SIRT1 is a posttranslational regulator of protein deacetylation and has been linked to a variety of pathological processes including inflammatory and stress responses [50]. *IMPA2* and *INPP1* are part of the phosphoinositol pathway, which has been linked to lithium’s mechanism of action, and genetic variants in these genes have been linked to suicide attempt in BD [5]. Because inflammation and lithium are both known to alter DNA methylation, we could hypothesize that genetic variation in these genes may predispose one for altered DNA methylation in the presence of certain environmental factors such as inflammation or medications. The molecular mechanisms underlying *cis*-meQTLs have not been established but there are some hypotheses. The passive hypothesis suggests that the inhibition of protein binding activity is enough to allow DNA methylation changes. The active hypothesis suggests that the inhibition of protein binding activity causes recruitment of DNMT or TET enzymes to modify the methylation status of nearby CpG sites [51]. Functional experiments are required to understand the biological mechanism and impact of the identified meQTLs.

Notably, there is very little overlap between our significant methylation results and the identified meQTLs. The findings that did overlap, did not survive correction for multiple testing in the initial analyses. These disparate results suggest that these meQTLs may not be related to suicide, and that the genetic variants and CpG sites chosen for this study are not *cis*-meQTLs. However, this does not rule out the possibility of meQTLs outside our regions of interest impacting our chosen CpG sites. Specifically, our *cis*-meQTL range of 15kb is stringent, with other definitions considering sites 1Mb away or on the same chromosome as a *cis*-meQTL [51]. To increase the power of our study, we did not investigate *trans-*meQTLs, which may also impact DNA methylation in our target regions.

This study has other limitations. Although our literature review was robust, most of these genetic regions have not been replicated and therefore may not represent truly suicide-related genomic loci. In addition, because this was a hypothesis-driven study, we purposely focused on only BD. Therefore, the results may not translate across psychiatric disorders. Finally, it is important to note that a history of suicide attempts, including number and lethality, do not consider time since attempt, which could introduce variability and reduce power. Additional considerations that could not be included were current medication and early childhood assessments, which have been shown to impact DNA methylation.

In summary, the current study illustrates that DNA methylation is altered in individuals with BD and a history of suicide attempt. The impacted genes play a role in immune response, a common theory of the pathophysiology of suicide. Additionally, the presence of meQTLs suggests that genetic variants influence the propensity for specific DNA methylation patterns in regions previously associated with suicidal behavior. Overall, there is substantial value to including meQTLs in future large scale suicide studies in BD.

## Data Availability

All data produced in the present study are available upon reasonable request to the authors.

## Acknowledgments

We would like to thank the study participants for their willingness to participate in the study. We would also like to thank Hsiang Wen and Benney Argue for sample collection and management of the cohort.

## Author Contributions

Design and conceptualization of the study: AA and MEG. Sample collection and recruitment: JGR, AW, JAW, VAM, and JGF. Data generation, processing, and statistical analyses: AA, LGC, and JM. Scientific discussion and interpretation of results: AA, LGC, AW, JAW, VAM, JGF, JM, and MEG. Wrote the manuscript: AA and MEG. All authors edited the manuscript.

