## Supplemental Tables 1, 2, and 3 for "Investigating the relationship between DNA methylation, genetic variation, and suicide attempt in bipolar disorder"

**Supplemental Table 1. Demographic Information**

| | No history of suicide attempt (N=51) | History of suicide attempt (N=55) | Overall (N=106) | P-value |
|---|---|---|---|---|
| ***Sex*** | | | | 0.132[a] |
| **Male** | 22 (43%) | 15 (27%) | 37 (35%) | |
| **Female** | 29 (57%) | 40 (73%) | 69 (65%) | |
| ***Age (years)*** | | | | 0.308[b] |
| **Mean (SD)** | 39.4 (13.7) | 42.1 (13.6) | 40.8 (13.6) | |
| **Median [Min, Max]** | 37.0 [18.0, 66.0] | 41.0 [18.0, 68.0] | 41.0 [18.0, 68.0] | |
| ***Race*** | | | | 0.404[c] |
| **Am. Indian/Alaskan Native** | 1 (2%) | 1 (2%) | 2 (2%) | |
| **Asian** | 1 (2%) | 0 (0%) | 1 (1%) | |
| **Black/African American** | 1 (2%) | 5 (9%) | 6 (6%) | |
| **More than one race** | 1 (2%) | 2 (4%) | 3 (3%) | |
| **Unknown** | 0 (0%) | 0 (0%) | 0 (0%) | |
| **White** | 47 (92%) | 47 (86%) | 94 (89%) | |
| ***Smoking history*** | | | | **0.024[c]** |
| **Current smoker** | 5 (10%) | 14 (26%) | 19 (18%) | |
| **Missing** | 0 (0%) | 1 (2%) | 1 (1%) | |
| **Never smoker** | 28 (55%) | 17 (31%) | 45 (43%) | |
| **Past smoker** | 18 (35%) | 23 (42%) | 41 (39%) | |
| ***Body Mass Index (BMI)*** | | | | 0.605[b] |
| **Mean (SD)** | 30.3 (7.61) | 31.0 (7.39) | 30.7 (7.47) | |
| **Median [Min, Max]** | 29.2 [18.4, 59.0] | 31.0 [18.9, 50.5] | 29.9 [18.4, 59.0] | |
| ***Number of lifetime suicide attempts*** | | | | **$1.831 \times 10^{-6}$[b]** |
| **Mean (SD)** | 0 (0) | 3.56 (4.94) | 1.85 (3.97) | |
| **Median [Min, Max]** | 0 [0, 0] | 2.00 [1.00, 30.0] | 1.00 [0, 30.0] | |

Descriptive statistics for the bipolar disorder cohort separated by history of suicide attempt. [a]Pearson's Chi-squared test with Yates' continuity correction; [b]Welch's Two Sample T-Test; [c]Fisher's Exact Test. Significant differences are indicated in bold. SD, standard deviation.

**Supplemental Table 2. Correlation between DNA methylation and number of lifetime suicide attempts**

| Gene | CpG Genomic Location | Statistic | P-value | False Discovery Rate (FDR) |
|---|---|---|---|---|
| MIF | chr22:23894125 | -3.664 | $2.49x10^{-4}$ | 0.022 |
| IMPA2 | chr18:11994457 | -3.136 | $1.71x10^{-3}$ | 0.075 |
| INPP1 | chr2:190354545 | -2.806 | $5.02x10^{-3}$ | 0.147 |
| IRX2* | chr5:2468211 | -2.645 | $8.17x10^{-3}$ | 0.177 |
| NOS3 | chr7:150998982 | -2.573 | 0.010 | 0.177 |
| CRHR1 | chr17:45805706 | -2.395 | 0.017 | 0.195 |
| MIF | chr22:23894167 | 2.287 | 0.022 | 0.195 |
| MIF | chr22:23894177 | 2.323 | 0.020 | 0.195 |
| IRX2* | chr5:2468198 | -2.341 | 0.019 | 0.195 |
| TGFBR1 | chr9:99146591 | -2.430 | 0.015 | 0.195 |
| ACP1 | chr2:273567 | 2.198 | 0.028 | 0.224 |
| INPP1 | chr2:190354502 | -2.145 | 0.032 | 0.235 |
| OXTR | chr3:8762680 | -2.055 | 0.040 | 0.270 |
| ANK3 | chr10:60124978 | -1.948 | 0.051 | 0.283 |
| BDNF | chr11:27658429 | -1.984 | 0.047 | 0.283 |
| INPP1 | chr2:190354478 | -1.966 | 0.049 | 0.283 |

Results with an uncorrected P-value less than 0.05 shown. GRCh38/hg38 used to annotate genomic locations. *Closest gene if variant is intergenic.

**Supplemental Table 3. Correlation between DNA methylation and severity of most lethal attempt**

| Gene | CpG Genomic Location | Statistic | P-value | False Discovery Rate (FDR) |
|---|---|---|---|---|
| CACNA1C | chr12:2613346 | -3.459 | $5.43x10^{-4}$ | 0.048 |
| CACNA1C | chr12:2613360 | -2.912 | $3.60x10^{-3}$ | 0.158 |
| BDNF | chr11:27658332 | -2.612 | $9.01x10^{-3}$ | 0.264 |
| ACP1 | chr2:273531 | 2.382 | 0.017 | 0.318 |
| OXTR | chr3:8762680 | -2.365 | 0.018 | 0.318 |
| IMPA2 | chr18:11994474 | -2.118 | 0.034 | 0.502 |
| IMPA2 | chr18:11994457 | -2.021 | 0.043 | 0.545 |

Results with an uncorrected P-value less than 0.05 are shown. GRCh38/hg38 used to annotate genomic locations.
